# Comparisons of Auditory Steady State and Auditory Brainstem Response Thresholds in Infants with Normal Hearing and Conductive Hearing Loss

**DOI:** 10.1101/2023.08.12.23294029

**Authors:** Hope Valeriote, Susan A. Small

## Abstract

This study investigates how well the air- and bone-conduction auditory steady-state response detects mild conductive hearing loss compared to the auditory brainstem response in young infants. Air-bone gap sizes are compared between infants with normal hearing and conductive loss using a two-group cross-sectional design. Twenty-three (500 Hz) and 22 (2000 Hz) infants (0-6 months of age) with normal hearing and 15 (500 Hz) infants with conductive loss were recruited from newborn hearing screening. Thresholds were obtained to frequency-specific air- and bone-conducted stimuli. There were no instances of conductive loss at 2000 Hz. 500 Hz mean thresholds and air-bone gap sizes were compared. Sensitivity and specificity for identifying conductive loss were measured. Overall, mean bone-conduction thresholds were similar between groups, and mean 500-Hz air conduction thresholds were higher with larger air-bone gap size for infants with conductive loss. Sensitivity and specificity for identifying conductive loss was highest for air-conduction auditory brainstem response threshold measurement compared to screening and auditory steady-state response threshold measurements. Compared to the auditory brainstem response, the variability of auditory steady-state response thresholds and air-bone gap size was too great to reliably separate normal hearing from mild conductive loss. More research is needed using infants with varying degrees of hearing loss at multiple frequencies to fully assess the appropriateness of the auditory steady-state response as a clinical diagnostic tool for an infant population.

---

Early hearing detection and intervention (EHDI) programs continue to be implemented internationally and are a major driving force for current research on infant hearing. Within this context, finding more efficient, less time-consuming methods of evaluating hearing in young infants has been of interest with some looking for alternatives to the current diagnostic gold-standard method, the auditory brainstem response (ABR), to integrate in their programs. For decades, the ABR has been the proven and reliable method that major EHDI programs (Bagatto, 2020; Hatton, Van Maanen & Stapells, 2022; JCIH, 2019) rely on to identify hearing loss in the infant population. Like behavioural methods of auditory assessment, ABR thresholds can be obtained using both air- and bone-conducted stimuli (Stapells & Ruben, 1989; Yang et al., 1987, 1993). Bone-conduction (BC) ABR assessment of infant hearing is required to differentiate between conductive and sensorineural hearing losses when air-conduction (AC) thresholds are elevated (Hatton et al., 2012). When conductive hearing loss (CHL) is present, an air-bone gap can be seen between elevated AC and normal BC thresholds.

Previous research to assess optimal testing conditions, including bone oscillator placement on the infant head and coupling method (i.e., handheld vs. band) and force, have informed clinical best practice in assessing BC thresholds in this young population who is unable to respond behaviourally (Small et al., 2007; Yang et al., 1991). The ASSR is another auditory evoked potential that is of interest to clinicians and researchers as an alternative to ABR because it also assesses hearing using both AC and BC stimuli, with the added benefit of testing multiple frequencies and ears simultaneously.The decision to use the ASSR as a method to assess infant hearing requires clear demonstration that the ASSR is comparable (or superior) to current gold-standards in its ability to assess infant AC and BC frequency-specific hearing thresholds in an accurate and precise way. An EHDI program requires high sensitivity to detect hearing loss in its target population and high specificity in differentiating elevated from normal hearing levels. Comparisons are needed between the ASSR and current gold-standard methods that take into account the participants’ age (i.e., behavioural audiometry as the gold standard for infants >6 months and ABR for infants ≤6 months (Gorga et al., 2006; JCIH, 2019; Widen et al., 2005). AC and BC modes of presentation, and hearing presentations from normal hearing ability to all degrees of sensorineural, conductive and mixed hearing loss. Maturational changes in BC ASSR responses and infant/adult differences in skull properties are discussed in Small & Stapells (2008a). Given these differences, it is important to understand how the BC ASSR behaves in infants with hearing loss. External ear canal changes and maturation for AC are understood and can be measured, but maturation effects for these infants are not as well documented for BC results. Understanding this is relevant to the use of ASSR/ABR in the clinical setting.

A review of the literature shows that many of these comparisons have been investigated and published over the last few decades and comparisons between ASSR and behavioural thresholds to date have been encouraging. For example, several studies have shown that AC and BC ASSR and behavioural thresholds in infants with normal hearing (NH) (Casey & Small, 2014; Luts et al., 2006) and infants with hearing loss (Aimoni et al., 2018) correlate highly. One study with a small number of participants also suggests the BC ASSR is able to identify normal cochlear sensitivity in young children with conductive hearing loss (Nagashima et al., 2013).

It is known that there are differences in BC ASSR thresholds by age likely due to skull maturation (discussed in detail in Casey & Small, 2014 and Small & Stapells, 2008a); however, the ASSR introduces other possible factors to consider, such as the use of high stimulus rates and multiple stimuli, that may have an effect on ASSR thresholds that does not apply to the ABR. It is especially important to establish a comprehensive body of literature for infants who are too young to respond behaviourally and who define the target population for EHDI programs. Studies comparing AC ASSR thresholds to the tone-ABR (using varied stimulus parameters and test protocols) thus far have shown that AC ASSR thresholds (in dB HL) in infants with NH and with hearing loss are consistently poorer than AC ABR thresholds (in dB nHL) but are highly correlated and accurate (Michel & Jørgensen, 2017; Rance et al., 2006; Rance & Rickards, 2002; Rodrigues et al., 2010; Van Maanen & Stapells, 2009, 2010). More recently, however, at least one other study using different collection protocols and stimulus parameters have suggested the reverse, with thresholds (in dB eHL) for ABR being poorer than for ASSR (Sininger et al., 2018) This latter study, however, used larger correction factors for ASSR compared to ABR, which may explain this finding. More studies with infants with hearing loss are needed, but the existing AC ASSR data appear promising. Studies that compare frequency-specific BC toneburst-ABR thresholds with sinusoidal amplitude modulated (SAM) tone ASSR thresholds in young infants with NH and hearing loss are significantly lacking, thus this is the focus on the present study. Small & Stapells (2008a) published BC ASSR normative data proposing a set of “normal” or minimum ASSR intensities, among others, for infants 0-11 months but did not compare frequency specific BC ASSR to BC ABR. Swanepoel et al. (2008) did provide some BC ASSR data for infants with hearing loss, but did not compare these results to toneburst ABR results and tested a broad age range (0.25-11.5 years of age). To our knowledge, no data comparing BC ASSR to BC ABR thresholds in young infants with hearing loss exist in the form of peer-reviewed publications.

For the present study, AC and BC ABR and ASSR thresholds in NH infants and infants with CHL confirmed by the gold-standard ABR thresholds were compared to investigate the following questions:

1. What are ABR and ASSR thresholds in young infants with NH and CHL?
2. How does the air-bone gap (ABG) compare between ABR and ASSR in young infants with NH and CHL?
3. What are appropriate minimum intensity cutoffs to differentiate NH from CHL using AC and BC ASSR in young infants?
4. Does the ASSR detect CHL as well as the ABR in young infants?

## Methods

Stimulus and recording setups for ABR followed those described in the BCEHP ABR protocol (Hatton et al., 2022). Stimulus and recording setups for ASSR were similar to those described in Casey & Small (2014), with minor differences between the research and clinical versions of the MASTER software. Details of methodology for ABR and ASSR are provided below.

### Participants

Infants were recruited through the newborn hearing screening program at the Royal University Hospital, Saskatoon. Participants were recruited if they failed newborn hearing screening or if they were unable to be screened at birth. Participation was entirely voluntary. Sixty four infants between the ages of 0 and 6 months participated (NH mean age = 7.36 weeks, range 0.6-12.9 weeks, CHL mean age = 6.71 weeks, range 2.9-20.6 weeks); 61 from the well-baby nursery and 3 graduates from the neonatal intensive care unit (NICU), none of whom presented with congenital aural atresia or microtia. Fourteen infants were excluded because they did not sleep and did not complete any conditions of the testing session.

Each frequency (500 and 2000 Hz) was assessed individually and was categorized as a normal hearing or conductive hearing loss threshold based on the relationship between the AC and BC ABR results. In other words, if AC ABR was within normal limits at a specific frequency, that frequency’s threshold was placed in the normal hearing group. If AC ABR was elevated with normal BC ABR results at the same frequency, the frequency’s threshold was placed in the conductive hearing loss group. Normal versus elevated levels correspond to those specified in the British Columbia Early Hearing Program (BCEHP) protocols. The BCEHP minimum stimulus intensities for 500 Hz are AC: 35 dB nHL BC: 20 dB nHL and for 2000 Hz are AC: 30 dB nHL and BC: 30 dB nHL (Hatton et al., 2022).

Results were included in the analysis whether partial or complete conditions were obtained. Infants who did not complete any portion of the protocol due to inability to sleep were excluded.

To verify the status of the middle ear and hearing at the time of testing, 1000 Hz tympanometry and transient-evoked otoacoustic emissions (TEOAEs) were performed using a Madsen AccuScreen and the OTOflex. The primary purpose of the screening measures was to corroborate the presence of middle ear pathology when participants were identified with CHL shown by abnormal tympanograms and absent TEOAEs and to determine the follow-up protocol as per the Royal University Hospital guidelines. The cross-check principle has been used in pediatric audiology for decades. As Hall (2016) describes, “no auditory test result should be accepted and used in the diagnosis of hearing loss until it is confirmed or crosschecked by one or more independent measures.” TEOAE stimulus levels ranged from 70-84 dB SPL and used noise-weighted averaging. The response detection method involved the counting of significant signal peaks with self-calibration depending on ear canal volume. To pass the TEOAE test, a total of eight valid peaks in alternating directions (counted both above and below the median line) must be present. Of the 50 participants who completed the testing, 31 did not pass tympanometry and 31 did not pass OAEs. A tympanogram was considered to be a “refer” if there was no identifiable peak, or maximum admittance was less than or equal to 0.6 mmho compensated from the negative tail at -400 daPa, and thus, the tympanogram was considered flat (type B). Type B tympanograms (Jerger, 1970) show minimal or no mobility of the tympanic membrane supportive of otitis media with effusion (OME) and is considered to be abnormal tympanometric pattern. TEOAEs were considered to be a “refer” if there was a response in fewer than three bands. Data collection took place in the context of a single audiology visit per participant, and the results of any medical and/or audiological follow-up is unknown.

### Stimuli

AC stimuli were presented to participants using an ER-3A insert earphones in one ear (the same ear that was used to establish BC thresholds). BC stimuli were presented to participants using the B-71 bone oscillator placed on the mastoid, slightly posterior to the upper portion of the pinna for both ABR and ASSR testing. Small et al. (2007) showed no difference between lower and upper mastoid bone oscillator placement, so the upper portion was chosen to avoid interfering with the nearby mastoid electrode. This was coupled to the head with approximately 400 grams of force using the hand-held method (i.e., held by the first author). This coupling method was used as it was the least disruptive method to the infants’ sleep and was found to have no significant differences to thresholds obtained by the elastic headband coupling method (Small et al, 2007). The examiner was trained to apply 400 grams of force (425 ± 25 g) by practicing BC application on a compressive spring scale, pressing down on the transducer with one or two fingers until the desired force was achieved with feedback. Once trials were completed with feedback, additional trials were completed without feedback, in a method similar to Small et al. (2007). Once it was determined that examiner was adequately trained, data collection began. Force was not verified on the infant head during testing. For ABR testing, the Intelligent Hearing Systems (IHS) SmartEP was used to generate and present stimuli. BCEHP-specified stimuli were used for both 500 and 2000 Hz (Hatton, Van Maanen, & Stapells, 2022). These stimuli were exact-Blackman-windowed tones (five-cycle total duration, no plateau) and presented at a rate of 39.1/sec to one ear (Hatton et al., 2019, 2022; Janssen et al., 2010). For ASSR testing, the two-channel Master II Clinical System was used to generate and present ASSR stimuli with carrier frequencies 500, 1000, 2000 and 4000 Hz. These stimuli were AM^2^ at modulation frequencies 78, 85, 93 and 101 Hz for carrier frequencies 500, 1000, 2000 and 4000 Hz, respectively, and were presented simultaneously to one ear (“monotic multiple, MM” ASSR).

### Calibration

#### ABR Stimuli

AC stimuli were calibrated in dB nHL using ppeSPL with a Quest 177 sound level meter and G.R.A.S. DB 0138 2-CC coupler with 1-inch microphone. The acoustic calibrations for 0 dB nHL for AC using insert earphones, were 22 and 20 dB ppe SPL for 500 Hz and 2000 Hz, respectively. BC stimuli were calibrated using the B & K 4930 artificial mastoid, where the acoustic calibration for 0 dB nHL for BC using the B-71 bone oscillator were 67 and 49 dB re: 1μN ppe at 500 and 2000 Hz, respectively [see Stapells & Small (2017), British Columbia Early Hearing Program (Hatton, Van Maanen, & Stapells, 2022) or Ontario Infant Hearing Program (Bagatto, 2020) protocols for Canadian ppeRETSPLs and ppeRETFLs].

#### ASSR Stimuli

AC ASSR stimuli were calibrated in ppe SPL using the dB SPL RETSPLs as per ANSI (1996) using a Quest 177 sound level meter and G.R.A.S. DB 0138 2-CC coupler with 1-inch microphone. Each of the four frequencies were calibrated separately in dB HL and then combined. Calibrations for 0 dB HL for AC using insert earphones was 5.5 and 3 dB SPL for 500 Hz and 2000 Hz, respectively (ANSI, 1996). BC stimuli were similarly calibrated using (ANSI, 1996) RETFLs with the Quest 177 sound level meter and B & K Mastoid 4930 artificial mastoid. Calibrations for 0 dB HL were 58 and 31 dB re: 1μN for BC at 500 and 2000 Hz, respectively.

### Recording

All participants were tested at the Royal University Hospital, Saskatoon in a double-walled sound-attenuating booth. All recordings were obtained using the Intelligent Hearing Systems SmartEP ABR system and the Master II Natus/Biologic clinical ASSR System. Four disposable electrodes were placed on the infant’s scalp using the typical electrode montage for infant ABR testing: one (non-inverting) electrode on the vertex, an (inverting) electrode on each mastoid and the common electrode off-center on the forehead. Impedance for each electrode was less than 3 kOhms.

For ABR testing, standard BCEHP parameters were used at the time of data collection. Gain was set to 100,000 and band-pass filtering from 30 to 1500 Hz with an artifact rejection of ±25 μV. One channel was recorded for AC ABR conditions and two channels were recorded (i.e., the ipsilateral and contralateral montages) for BC ABR conditions. A minimum of two replications of 2000 trials each was obtained at threshold levels and one step (of 10 dB) below threshold. The presence and/or absence of a response were determined visually and by objective measures (signal to noise ratio and residual noise) and was interpreted by the first author. A response being “present” was determined by a visually identifiable wave V in the averaged waveform in the ipsilateral channel recording for AC ABR and by an ipsilaterally dominant wave V response for BC ABR when ipsilateral and contralateral recordings were compared (Hatton et al., 2022). In accordance with BCEHP guidelines, “no response” was determined only when no visually identifiable wave V was present and SmartEP residual noise was less than or equal to 0.08 μV (Hatton, Van Maanen, & Stapells, 2022). For ABR measures, where a response was identified, RN and SNR measures were used to support visual interpretation where possible, but ultimately visual identification of a response was considered sufficient to determine “response present”. “No response” judgements were made on the basis of both visual interpretation but also with IHS-SmartEP SNR values <1 and RN ≤0.08uV. The participants were classified as having CHL at a frequency by demonstrating elevated AC ABR results with normal BC ABR results (based on BCEHP levels) with abnormal tympanometry findings. In the classification of participants, tympanometry and OAE screening was used only as a cross-check to confirm CHL where identified at a specific frequency to provide additional evidence in the identification of CHL. The NH group did not have a specific criterion for tympanometry or OAE screening result and that categorization was made on the basis of present ABR to AC and BC stimuli at minimum normal levels at that frequency (Hatton et al., 2022). To ensure the validity in this method of categorization, an independent samples t-test was performed comparing AC ABR and ASSR NH thresholds in infants with normal OAE screening results and abnormal OAE screening results for 500 Hz and 2000 Hz separately and was not significant. This supports the validity of this method of categorization. Where the typical clinical protocol used in BCEHP does not include testing down to threshold if a present response has been established at the minimum stimulus intensity, testing down to threshold for all measures completed (AC and BC ABR and ASSR) did take place in this study.

For ASSR testing, two-channels were recorded but only the ipsilateral channel (i.e., vertex-ipsilateral mastoid) was examined when determining response presence or absence and was the only channel analyzed in this study. For ASSR measures, no visual identification was required, as only SNR (p value) and residual noise were used to make response/no-response determinations. Masking was not used as the interaural attenuation for infants reported by Small & Stapells (2008b) is at least 10-30 dB. The EEG was filtered using a 30-150 Hz filter and amplified 10 000 times with artifact rejection set to ±125 μV. The analog-to-digital conversion rate was 1200 Hz. Each sweep consisted of 16 epochs of 1024 data points and took 13.107 seconds of recording time. The ASSRs were averaged in the time domain and analyzed online in the frequency domain using a fast Fourier transform (FFT) with a resolution of 0.076 Hz over a range of 0-625 Hz. Amplitudes were measured baseline-to-peak and expressed in nV. Recording continued until there was a response present with a minimum of 10 sweeps, or the residual noise levels were at least <15 nV and there was a minimum of 10 sweeps completed; whichever came first. A F-ratio was calculated by the MASTER II system and a response was considered present if a significant response value (p<0.05), was obtained from the F-ratio compared to critical values for F(2, 240) for at least three consecutive sweeps. The F-ratio estimated the probability that the amplitude of the ASSR at the modulation frequency was significantly different from the average amplitude of the noise at adjacent frequencies. This was calculated within 120 bins, or +/-60 bins from the modulation frequency (John & Picton, 2000). A response was considered absent if no significant response value was obtained (p>0.05) and the noise value was appropriately low (<15 nV).

### Procedure

One session lasting between one and three hours took place for each subject. Before testing began, caregivers consented to participation in the study and were provided a small honorarium. All infants completed a hearing screening (tympanometry and transient evoked otoacoustic emissions [TEOAEs]), ABR and ASSR testing in the recording session. One ear was chosen to be tested using electrophysiologic methods. The selection of test ear was made based on the outcome of the hearing screening. If only one ear failed, that ear was tested using ABR and ASSR; if both ears failed, or both ears passed, the ear was chosen based on the most comfortable position for the infant and the caregiver. Testing was completed with the examiner inside the booth, next to the infant and caregiver. The examiner held the oscillator and continually monitored the placement of the earphone in the infant’s ear.

Hearing screening using TEOAEs and 1000 Hz tympanometry was conducted and electrodes were applied while the infant was awake. The infant was given the opportunity to fall asleep before ABR and ASSR testing began and remained asleep during these tests in the caregiver’s arms during ABR and ASSR testing. If the infant woke during the session, an opportunity for them to fall back of sleep was given before testing continued. Electrophysiological testing always began with ABR in order to provide parents with information from a “gold-standard” test before proceeding with ASSR. AC ABR was followed by BC ABR.

ABR testing began at BCEHP minimum stimulus intensities that correspond to the upper limit of normal hearing. For AC ABR, testing began at 35 dB nHL and 30 dB nHL for 500 Hz and 2000 Hz, respectively. BC ABR testing began at 20 dB nHL and 30 dB nHL for 500 and 2000 Hz, respectively. No masking was used given the age group of the participants and large interaural attenuation. A 10-dB bracketing method was used. Threshold levels were defined as the lowest level at which a response is present with an absent response 10 dB below. The lowest level tested was 0 dB nHL for 2000 Hz AC and BC and 500 Hz BC, and 5 dB nHL for 500 Hz AC due to starting levels and 10 dB step sizes.

ASSR testing began at 30 dB HL, corresponding to the highest “minimum level” obtained by Casey & Small (2014). Similar to the ABR testing procedure, a 10-dB bracketing method was used, and threshold was defined as the lowest level at which a response is present with an absent response 10 dB below. The lowest level tested was 0 dB HL for ASSR. As the study was more interested in BC comparisons, BC ASSR was prioritized over AC and was completed first. Thresholds were found using a 10-dB bracketing procedure. If a response was present, intensity was decreased by 10 dB. If no response was present, intensity was increased by 20 dB. Testing continued down to threshold.

In the sections to follow, only 500-Hz data for NH and CHL groups will be discussed. Although 2000-Hz data were collected (shown in Table 2), as there were no instances of CHL observed at 2000 Hz, these data are only briefly addressed in the remainder of this article.

### Data Analyses

#### AC and BC ABR and ASSR Thresholds

Measures of central tendency and dispersion are provided for each frequency condition (500 & 2000 Hz) by group (NH, CHL). Independent samples t-tests were performed to compare 500 Hz AC and BC ABR and ASSR thresholds between NH and CHL groups.

#### Air-Bone Gaps

Air-bone gap (ABG) between ABR and ASSR in infants with NH and CHL were compared. Air-bone gaps (ABG) were calculated for ABR by subtracting the BC (nHL) threshold from AC (nHL) threshold for each subject. ASSR ABGs were calculated by subtracting the BC (dB HL) threshold from the AC (dB HL) threshold. For 500 Hz, an independent samples t-test was then conducted to determine if means between groups were significantly different. Differences in thresholds were considered significant at the p<0.05 level.

#### Minimum ‘Normal’ Intensities

Individual and mean AC and BC ABR and ASSR thresholds were determined for NH (500 and 2000 Hz) and CHL (500 Hz) groups. “Minimum normal intensities” represent the minimum test intensity a response would need to be present in a clinical setting to confirm normal hearing. Minimum intensities were determined by calculating the cumulative percent of responses present at each stimulus level for each testing method and mode of presentation. The intensity at which greater than 90 percent of normal-hearing infants had a response was considered the “minimum intensity” (e.g., Van Maanen & Stapells, 2009; Small & Stapells, 2008a). Ninety percent was chosen to represent a intensity that separates “normal” from “elevated” well according to the gold-standard threshold measure, ABR. The intensities for BC ABR have been assessed by Hatton et al. (2012).

#### Sensitivity and Specificity for Conductive Hearing Loss Detection with ASSR

Sensitivity and specificity using ASSR was measured using the ABR as the gold standard to determine whether the ASSR detects CHL as well as the ABR. Sensitivity and specificity of the OAE/tympanometry screening, AC ASSR threshold and air-bone gap and ABR air-bone gap were also measured. These were calculated as shown in Table 1. The 500 Hz ABR and ASSR thresholds were averaged separately for AC and BC, and 500 Hz thresholds were compared between normal and CHL groups. Analyses were performed using an independent samples t-tests. Differences in thresholds were considered significant at the p<0.05 level.

**Table 1.**
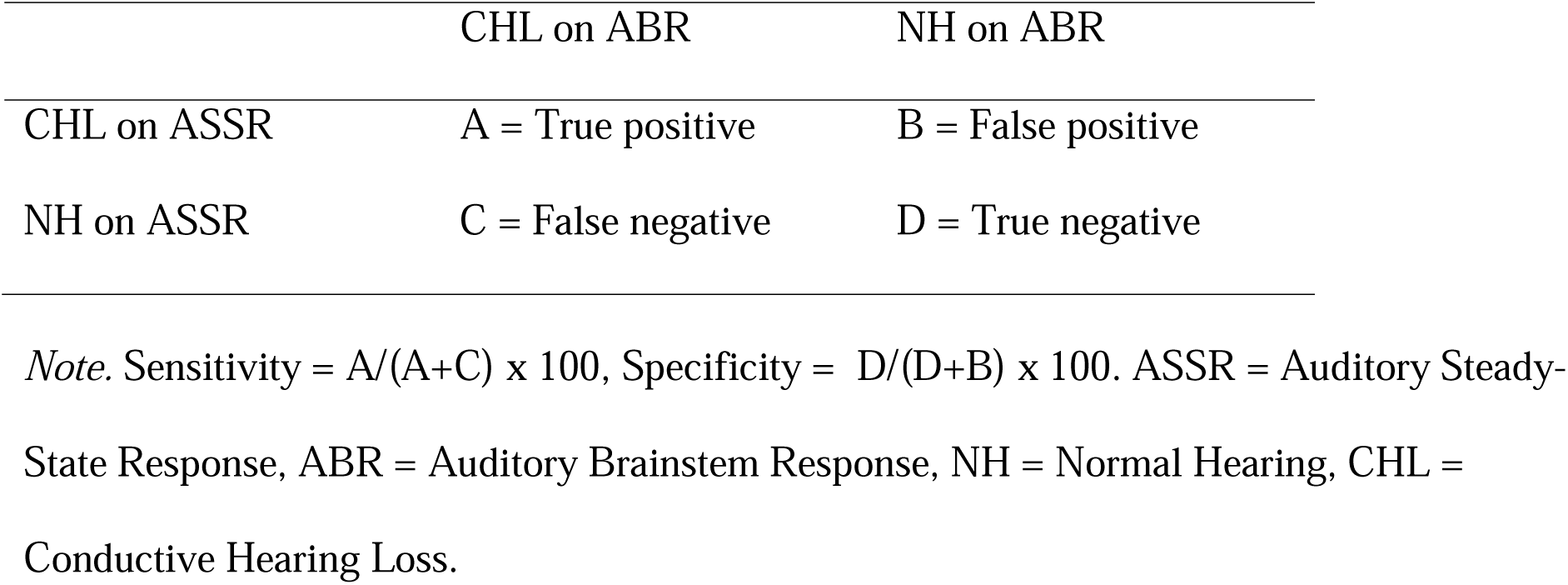
Sensitivity and Specificity Calculation Method.

|  | CHL on ABR | NH on ABR |
| --- | --- | --- |
| CHL on ASSR | A = True positive | B = False positive |
| NH on ASSR | C = False negative | D = True negative |

*Note.* Sensitivity = $A/(A+C) \times 100$ , Specificity = $D/(D+B) \times 100$ . ASSR = Auditory Steady-State Response, ABR = Auditory Brainstem Response, NH = Normal Hearing, CHL = Conductive Hearing Loss.

## Results

Mean thresholds (as well as SD and 90% levels) for both 500 and 2000 Hz ABR/ASSR AC and BC thresholds can be found in Table 2. For infants with confirmed CHL, mean AC ABR thresholds increased compared to infants with NH. Mean AC ABR and ASSR thresholds at 500 Hz were larger for infants with CHL. Standard deviations were greater for ASSR thresholds compared to ABR thresholds for both AC and BC. No infants demonstrated CHL at 2000 Hz (as defined as an elevated 2000 Hz AC threshold in the presence of a 2000 Hz BC threshold within normal limits) and for this reason are not discussed in sections to follow.

**Table 2.** AC and BC 2000 and 500 Hz ASSR and ABR mean thresholds for NH and CHL groups.

|  |  | NH Group |  |  |  | CHL Group |  |
| --- | --- | --- | --- | --- | --- | --- | --- |
|  |  | 2000 Hz |  | 500 Hz |  | 500 Hz |  |
|  |  | ASSR<br>(dB<br>HL) | ABR<br>(dB nHL) | ASSR<br>(dB HL) | ABR<br>(dB nHL) | ASSR<br>(dB HL) | ABR<br>(dB nHL) |
| AC | Mean | 20.47 | 17.72 | 29.52 | 25.43 | 36.67 | 48.33 |
|  | SD | 12.03 | 9.22 | 9.20 | 7.05 | 11.12 | 4.88 |
|  | n | 21 | 22 | 21 | 23 | 15 | 15 |
|  | 90% Level | 40 | 30 | 40 | 35 |  |  |
| BC | Mean | 21.00 | 15.00 | 17.39 | 10.41 | 15.33 | 12.00 |
|  | SD | 13.96 | 10.12 | 9.63 | 7.50 | 12.63 | 7.93 |
|  | n | 21 | 22 | 23 | 24 | 15 | 15 |
|  | 90% Level | 40 | 30 | 30 | 20 | 40 | 20 |

*Note.* AC = Air-Conduction, BC = Bone-Conduction, ASSR = Auditory Steady-State Response, ABR = Auditory Brainstem Response, NH = Normal Hearing, CHL = Conductive Hearing Loss, SD = Standard Deviation, n = number of participants, 90% Level = lowest level (in dB HL or nHL) at which at least 90% of group shows response present.

### 500 Hz ABR and ASSR Thresholds for CHL and NH Groups

Independent samples t-tests showed AC ABR thresholds for the CHL group were higher than thresholds for the normal-hearing group[t(36) = -10.95, p<0.001]. BC thresholds did not significantly differ across groups [t(37) = -0.67, p=0.51]. AC ASSR thresholds for the CHL group were also higher than thresholds for the NH group [t(34) = -2.10, p=0.043]. BC ASSR thresholds did not significantly differ across groups [t(36) = 0.56, p=0.579]. Levene’s Test for equality of variances was not significant, so equal variances were assumed. AC ABR and ASSR and BC ASSR and ABR thresholds were not correlated [AC slope = 0.17, intercept = 26.67, r^2^ = 0.04; BC slope = 0.15, intercept 15.35, r^2^ = 0.01].

### Air-bone Gap for CHL vs. NH Groups

The air-bone gaps (ABGs) at 500 Hz for the groups with NH and CHL are shown in Table 3, Figure 1 (ABR) and Figure 2 (ASSR). On average, the mean 500-Hz ABGs for the two groups were as follows: (i) ABR NH: 15 dB, (ii) ASSR NH: 12 dB (iii) ABR CHL: 36 dB, (iv) ASSR CHL: 21 dB. Two outliers were present in the ABR ABG data set for the CHL group. The independent samples bootstrapped t-test comparing mean ABR ABG for the NH and CHL group showed significantly larger ABGs for the CHL group than in the NH group [t(35) = -7.74, p<0.001]. Outliers were defined as a data point that is greater than the upper quartile + 1.5x the interquartile range. A bootstrapped t-test was used to account for the outliers. Similarly, for the ASSR, the independent samples t-test showed ABGs were also significantly larger in CHL group [t(33) = -2.30, p=0.028]. Levene’s Test for equality of variances was not significant, so equal variances were assumed. The majority of NH participants had 500-Hz ABR ABGs 25 dB or smaller (21 of 22) and CHL subjects of 35 dB or larger (13 of 15). As shown in Figures 1 and 2, compared to ABR, there was more overlap between ASSR NH and CHL groups where the majority of NH participants had ASSR ABGs of 20 dB or smaller and CHL of 10 dB or larger. ABR and ASSR ABG size, however, were poorly correlated (slope = 0.24, intercept = 10.02, r^2^ = 0.07). For 2000 Hz, the mean ABG for the NH group was approximately -3 and 3 dB for ASSR and ABR, respectively; the majority of NH infants had 2000-Hz ABR ABGs 10 dB or smaller (20 of 22), and 10 dB or smaller for ASSR (19 of 21).

**Figure 1.**
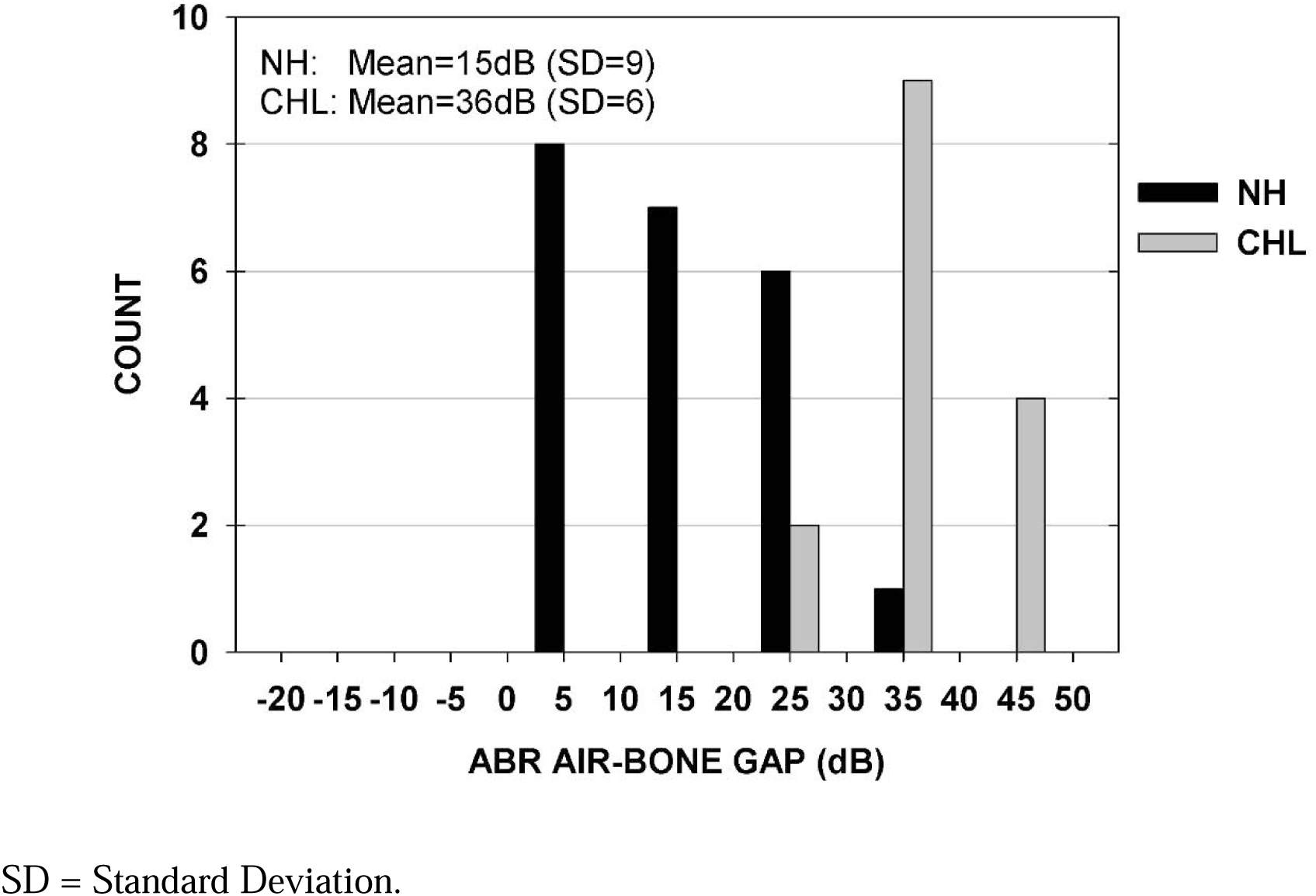
Auditory brainstem response (ABR) air-bone gaps (ABG) in normal hearing (NH) and conductive hearing loss (CHL) groups.

**Figure 2.**
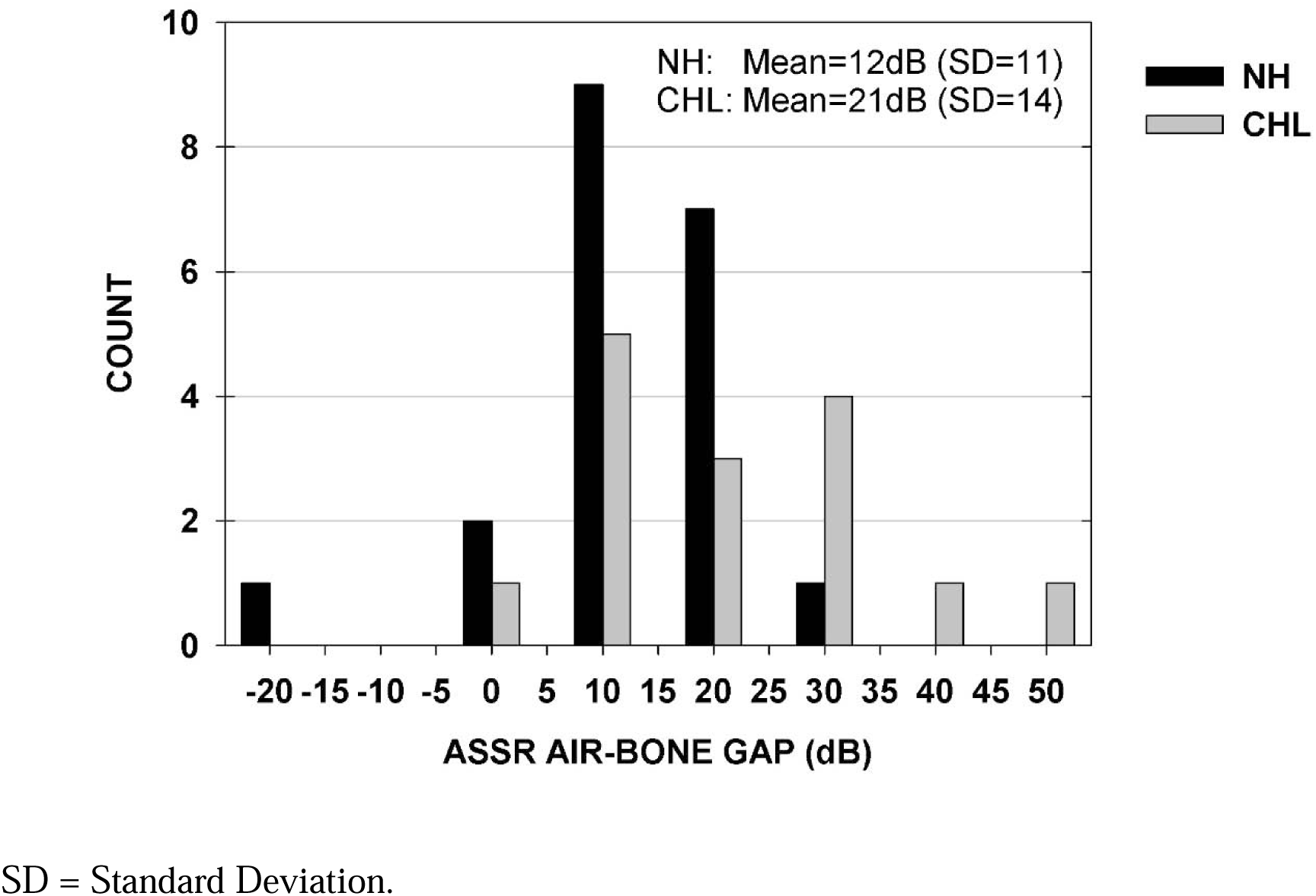
Auditory steady-state response (ASSR) air-bone gaps (ABG) in normal hearing (NH) and conductive hearing loss (CHL) groups

**Table 3.** 2000 and 500 Hz ASSR and ABR mean ABG size for NH and CHL groups.

|  | NH Group |  |  |  | CHL Group |  |
| --- | --- | --- | --- | --- | --- | --- |
|  | 2000 Hz |  | 500 Hz |  | 500 Hz |  |
|  | ASSR<br>ABG<br>(dB) | ABR<br>ABG<br>(dB) | ASSR<br>ABG<br>(dB) | ABR<br>ABG<br>(dB) | ASSR<br>ABG<br>(dB) | ABR<br>ABG<br>(dB) |
| Mean | -3.33 | 2.73 | 12.00 | 15.00 | 21.33 | 36.33 |
| SD | 15.92 | 12.41 | 10.56 | 9.26 | 13.56 | 6.40 |
| n | 21 | 22 | 20 | 22 | 15 | 15 |

*Note.* ASSR = Auditory Steady-State Response, ABR = Auditory Brainstem Response, ABG = Air-Bone Gap, NH = Normal Hearing, CHL = Conductive Hearing Loss, SD = Standard Deviation, n = number of participants.

### Minimum “Normal” Intensities

The minimum intensity cutoffs for ABR and ASSR were determined for AC and BC separately by calculating the cumulative percent of responses present at each stimulus intensity for each testing method and mode of presentation. The intensity at which greater than 90 percent of NH infants had a response was considered the “minimum intensity”. For ABR, the minimum intensity levels were 30 dB nHL for 2000 Hz AC and BC, and 35 and 20 dB nHL for 500 Hz AC and BC, respectively (see Table 2). For ASSR, the minimum intensity levels were 40 dB HL for 2000 Hz AC and BC and 40 and 30 dB HL for 500 Hz AC and BC, respectively. Minimum ASSR intensities (in dB HL) were found to be higher for AC compared to BC and higher for ASSR compared to the ABR minimum intensities (in dB nHL) used by the BCEHP.

### Sensitivity and Specificity for CHL Detection

Table 4 compares sensitivity and specificity for each test completed using different cutoff criteria. Sensitivity and specificity is calculated, with reference to NH or CHL diagnosis using the gold standard ABR, for: Tympanometry/OAE screening, ASSR and ABR ABG size, and AC ASSR thresholds. The subtype for each test category that had the highest specificity and a sensitivity exceeding 90% are bolded. For such a test to be used in the context of an early hearing detection and intervention program, there is very low tolerance for missing cases of hearing loss, so sensitivity must be high. For screening measures, sensitivity was high for all screening tests, while specificity was poor. For ABG, the sensitivity worsens and specificity improves as the criteria to define the minimum ABG for CHL increases. Using AC ASSR thresholds as the criterion for CHL identification, as threshold increases, sensitivity worsens while specificity improves.

**Table 4.** Sensitivity and Specificity of CHL detection compared to gold standard ABR by test type.

|  | Test | Sensitivity | Specificity |
| --- | --- | --- | --- |
| Screening | Tymp + OAE | 100* | 44 |
|  | OAE only | 100* | 58 |
|  | **Tymp only** | **100\*** | **60** |
| ASSR ABG size<br>(dB) | **$\geq 10$** | **93** | **15** |
| | $\geq 20$ | 60 | 60 |
| | $\geq 30$ | 40 | 95 |
| | $\geq 40$ | 13 | 100 |
| ABR ABG size<br>(dB) | $\geq 5$ | 100 | 0 |
| | $\geq 15$ | 100 | 36 |
|  | **$\geq 25$** | **100** | **68** |
| | $\geq 35$ | 87 | 95 |
| | $\geq 45$ | 26 | 100 |
| ASSR thresholds<br>(dB HL) | **AC $\geq 20$** | **100** | **5** |
| | AC $\geq 30$ | 87 | 33 |
| | AC $\geq 40$ | 47 | 67 |
| | AC $\geq 50$ | 33 | 100 |

*Note.* \*Sensitivity of 100% due to classification criteria for CHL that required “refer” results on both tympanometry and OAE screening. CHL = Conductive Hearing Loss, ABR = Auditory Brainstem Response, ASSR = Auditory Steady State Response, Tymp = Tympanometry, OAE = Otoacoustic Emissions, ABG = Air-Bone Gap, AC = Air-Conduction.

## Discussion

### ABR vs ASSR threshold differences

ABR and ASSR thresholds differed when measured in the same infant and were found to be poorly correlated in the present study (AC r^2^ = 0.04; BC r^2^ = 0.01). Given the narrow range of thresholds obtained, this is not surprising. Previous studies have shown strong correlations between ABR and ASSR thresholds, and they are thought to measure responses from approximately the same part of the auditory system (Van Maanen & Stapells, 2010; Sininger et al. 2018). For these reasons, it was expected that the ABR and ASSR thresholds would be similar. Threshold differences in the present study may be in part due to the 0 dB HL/nHL values used for ABR and ASSR, starting intensities being offset by 5 dB (i.e., ABR at 35 dB nHL and ASSR 30 dB HL), a 10-dB step size, and the rather modest AC threshold shifts due to mild CHL. If smaller step sizes, similar starting levels, and populations with greater degrees of CHL were used, this difference may have followed the trend of Rance et al. (2006) where the difference between thresholds became minimal when converted to like units. Other contributors to the ASSR-ABR differences are potentially related to an insufficiently large sample size and a difference in stimuli.

### Air-Bone Gaps

To our knowledge, this is the first study to compare AC and BC ASSR thresholds within participants in infants with ABR-confirmed NH and CHL. ABGs for both ABR and ASSR CHL groups were larger than their NH counterparts, as expected. We had anticipated that threshold differences would be observed between ASSR and ABR thresholds because dB nHL threshold values were used for ABR (i.e., thresholds that included a consideration of temporal integration issues) whereas the dB HL values for SAM tones were based on dB HL values for long-duration tones as specified in ANSI S3.6. However, we also anticipated the differences in threshold for ASSR and ABR to be similar for AC and BC stimuli, thus no impact to estimated ABGs was expected (i.e., whatever differences may be present between ABR and ASSR would affect AC and BC thresholds similarly and thus the ABG would not be greatly affected across these measures). ABR and ASSR ABGs for the NH group did not differ significantly, however CHL ABR and ASSR ABGs did differ significantly.

We also expected that the ABGs for ABR and ASSR would be larger in infants with CHL than with NH and this was confirmed. In clinical practice when using behavioural methods of assessment, clinicians operate under the assumption that individuals with NH and sensorineural hearing loss do not exhibit clinically significant ABGs; while those with CHL are expected to show an ABG. In adult audiometry, with test/retest reliability in behavioural audiometry of ±5 dB, an ABG ≥15 dB is often considered clinically significant and this tends to be extrapolated in clinical practice to define clinically significant ABG sizes in pediatric assessments.

In the clinical setting, it is challenging to assess the magnitude of the ABR and ASSR ABGs for several reasons. First, most ABR/ASSR clinical protocols do not encourage testing down to true threshold (at least, not in Canada), but rather recommend the use of “minimum normal intensities” where if a response is present, it is considered within normal limits and testing at lower presentation intensities is not required. The goal of most Canadian EHDI programs is to detect permanent congenital hearing losses ≥30 dB HL and this method of assessment accomplishes this goal in a time-effective manner. Second, ABGs for clinical use (in many provinces in Canada) are calculated on the estimated hearing levels (eHL) values for ABR after nHL-to-eHL correction factors have been applied. The purpose of these frequency- and mode-specific correction factors is to more closely estimate pure-tone behavioural (dB HL) thresholds used for diagnostic and hearing aid fitting purposes (see the most current BCEHP clinical protocol for up-to-date correction factors, corresponding stimulus parameters and recording techniques; Hatton et al., 2022). When AC and BC eHL correction factors are applied, they come with their own estimation errors (as much as 10-20 dB of error in either direction), and these errors are additive when calculating the ABG. These estimation errors in combination with using 10-dB step sizes and/or not testing down to a true threshold make ABG estimations in clinical practice challenging. The ABR or ASSR ABG can be substantially under- or over-estimated, and therefore are more appropriately used in a descriptive way to comment on the size of a conductive component rather than a singular diagnostic criterion. It is important that clinical protocols recognize these limitations. Clinicians need to keep in mind that unique correction factors are applied to each individual frequency and differ for AC and BC. When investigating the diagnostic power of ABG size, this can apply only to a specific frequency and cannot not be generalized beyond the frequency that is being investigated. Carefully determined correction factors are necessary and will affect any measure of the ABG. More data are needed for each frequency with different degrees of CHL. This study continues to support the value of the ABG as a descriptive tool to *accompany* frequency-specific ABR thresholds and tympanometry measures in differentiating NH from CHL.

### Normal Hearing Thresholds and Minimum Intensities

As shown in Tables 5 and 6, across several studies, normal ASSR intensity levels (i.e., ASSR threshold level upper limits for those infants with no hearing loss) are approximately 50 and 40 dB HL for 500 and 2000 Hz AC, and approximately 20-30 and 40 dB HL for BC stimuli. The present study showed minimum intensities of 40 and 40 dB HL for 500 & 2000 Hz AC and 30 & 40 dB HL for 500 & 2000 Hz BC for ASSR. The AC values differ by not more than 10 dB from the average of the other studies using different stimuli (note variations across studies in the modulation function, or use of one versus multiple simultaneous ASSR stimuli). These differences may be attributed to differences in sample size, stimuli, whether presentation was multiple or single, the age range tested, stopping criteria, EEG noise, recording system (and the system’s detection algorithm). The BC minimum intensity at 500 Hz proposed in Small & Stapells (2008b) of 30 dB HL is consistent with what was found in the present study. The minimum intensity from this study was slightly higher than 20 dB HL reported by Casey & Small (2014), however, was likely somewhat overestimated (∼5 dB) due to the distribution of the threshold data and the step size used. The present study and that by Casey & Small (2014) are the only studies to date that used AM^2^ stimuli for BC ASSR, and both studies showed better than the average of thresholds included in the table. However, ASSR amplitudes are in keeping with these studies and a study using AM/FM stimuli (Small & Stapells, 2008a). The mechanisms underlying this difference in thresholds remains an open question. In addition, maturation of the BC ASSR response makes “cutoff” points less clear for categorizing hearing loss and the use of an age range of 0-6 months may be too large. Minimum intensities for ABR were in keeping with those suggested by BCEHP. Importantly, they were not found to be lower. As mentioned earlier, t-tests comparing NH participants’ AC and BC ABR or ASSR thresholds at 500 and 2000 Hz between those with normal and those with abnormal OAE screening results showed no significant difference; thus, the NH group includes all NH thresholds, regardless of screening result. Minimum normal intensities for ABR also did not differ between those in the NH group with a normal vs. abnormal OAE screen; both subgroups had minimum normal intensities of 30 for 2000 Hz AC and BC and 35 and 20 for 500 Hz AC and BC, respectively.

**Table 5.** A summary of 500 & 2000 Hz AC ASSR “normal levels” in young children in literature.

|  | Modulation | Multiple (M)<br>/Single (S) | Age | Norm Max (dB HL) |  |
| --- | --- | --- | --- | --- | --- |
|  |  |  |  | 500 Hz | 2000 Hz |
| Lins et al., (1996) ** | AM | M | 1-10 mos | 48 | 38 |
| Cone-Wesson, Parker, et al., (2002) | AM | S | <4 mos | >71 | 50 |
| John, Brown et al., (2004) | MM, AM, AM<sup>2</sup> | M | 3-15 wks | >46 | >50 |
| Rance et al., (2005) | MM | S | 1-3 mos | 52 | 40 |
| Swanepoel & Steyn (2005) | MM | M | 3-8 wks | 50 | >50 |
| Luts et al., (2006) ** | MM | M | <3 mos | >44 | 42 |
| Rance & Tomlin (2006) | MM | S | 6 wks | 50 |  |
| Van Maanen & Stapells (2009) | AM (cos<sup>3</sup> sinusoids*) | M | <6 mos | 49 | 36 |
| Casey & Small (2014) | AM<sup>2</sup> | M | 6.5-19 mos | 30 |  |
| Rodrigues & Lewis (2014) ** | NBchirps | M | 2 days | 59 | 31 |
| Present Study | AM<sup>2</sup> | M | 0-6 mos | 40 | 40 |

*Note.* \*Cosine3-windowed sinusoids are nearly equivalent to AM<sup>2</sup>. \*\* Thresholds were converted from dB SPL to dB HL using ANSI-1996 adjustment values. \*\*\*Levels provided are in dB HL, converted levels from NB chirp « nHL » using Haughton 2-cc coupler conversions. AC = Air-Conduction, ASSR = Auditory-Steady State Response, AM = Amplitude Modulation, MM = Mixed Modulation, NB = Narrowband, Mos = Months, Wks = Weeks.

**Table 6.**
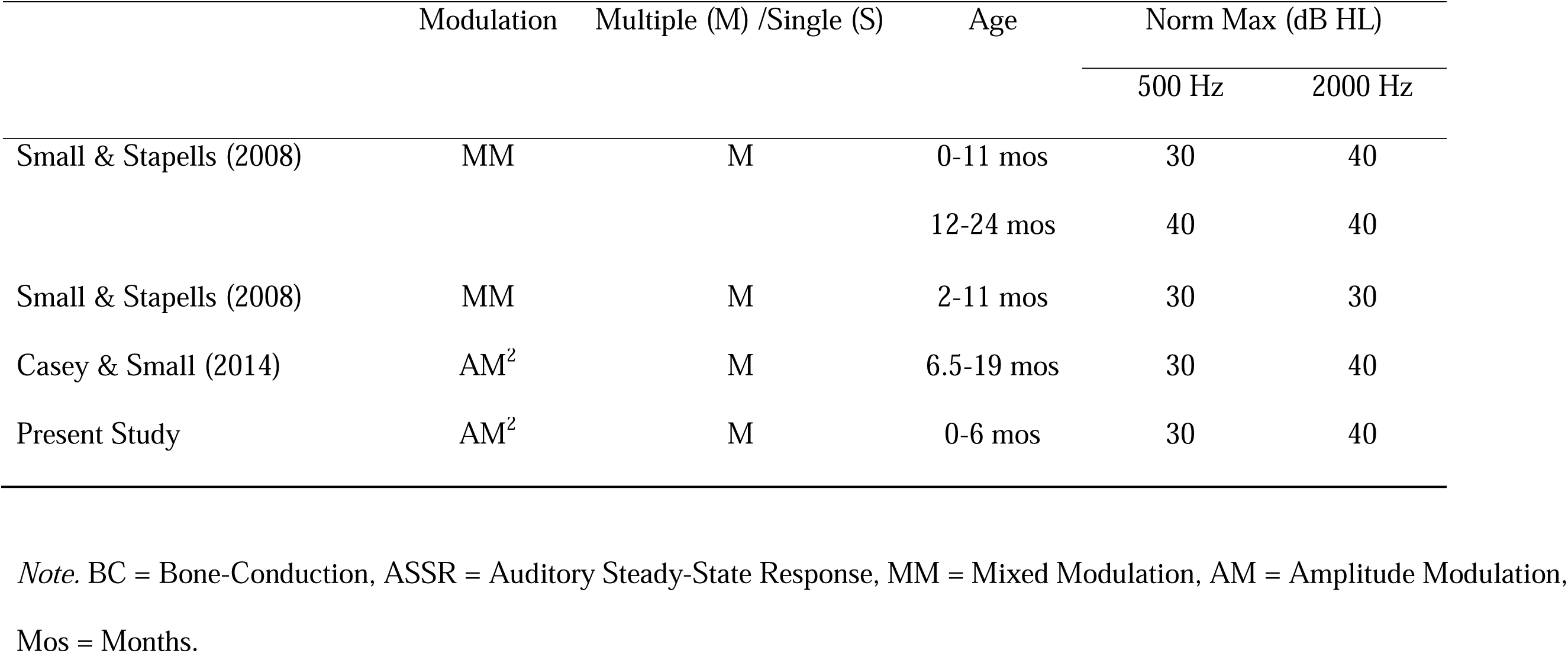
A summary of 500 & 2000 Hz BC ASSR “normal levels” in young children in literature.

|  | Modulation | Multiple (M) /Single (S) | Age | Norm Max (dB HL) |  |
| --- | --- | --- | --- | --- | --- |
|  |  |  |  | 500 Hz | 2000 Hz |
| Small & Stapells (2008) | MM | M | 0-11 mos | 30 | 40 |
|  |  |  | 12-24 mos | 40 | 40 |
| Small & Stapells (2008) | MM | M | 2-11 mos | 30 | 30 |
| Casey & Small (2014) | AM<sup>2</sup> | M | 6.5-19 mos | 30 | 40 |
| Present Study | AM<sup>2</sup> | M | 0-6 mos | 30 | 40 |

*Note.* BC = Bone-Conduction, ASSR = Auditory Steady-State Response, MM = Mixed Modulation, AM = Amplitude Modulation, Mos = Months.

### Sensitivity and Specificity for Conductive Hearing Loss Detection

As discussed earlier, 500-Hz AC ASSR thresholds are, on average, higher than BC ASSR thresholds in the CHL group, with a significantly larger ABG compared to the NH group (see Tables 2 and 3). In the case of mild CHL, however, the ABR does seem better at identifying modest ABGs in infants than the ASSR, using the parameters incorporated in this study. For this reason, clinicians should exercise caution when considering using ASSR AC/BC thresholds to identify mild CHL with small ABGs. Cutoff thresholds where the ASSR ABG demonstrated high sensitivity lacked high specificity (See Table 4). The sample for the current study did not include infants with sensorineural hearing loss, and therefore it was not possible to determine the number of false negatives for BC ASSR. There was significant overlap in the NH and CHL AC ASSR threshold distributions (and hence the imperfect sensitivity and specificity). We hypothesize that this overlap may be a reflection of the sampling between groups. Overall, the CHL group only demonstrated a very mild degree of hearing loss that was isolated to 500 Hz (and perhaps in some cases may have been resolving). The NH sample recruited was primarily “at risk” infants who failed or missed their initial hearing screening, and thus some subclinical degree of middle ear dysfunction may have been present. Perhaps if the CHL group demonstrated elevated AC thresholds across frequencies or the degree of loss was greater, and the NH group had instead been recruited from low-risk infants with confirmed normal middle-ear function, the overlap would have been minimized. Nevertheless, the present study operated under conditions that are typical in the clinical setting where the separation between these groups may be less than ideal.

## Clinical Implications

Previous studies that compared ASSR thresholds to gold-standard methods of infant hearing assessment reported strong correlations between the methods and reasonable accuracy in estimating hearing thresholds. Some studies have suggested that the ASSR can accurately separate NH infants from those with hearing loss. Differing methodology in this body of research continues to be problematic. This study is one of only a small number of studies comparing the ASSR to tone-ABR in infants with NH and with hearing loss, and within these, methodologies and stimulus choices differ. More work still needs to be done to determine the best methodology and stimulus parameters for ASSR testing in infants. This study added to the body of research by providing data using AC and BC ASSR to AM^2^ tones at 500 Hz in infants with NH or conductive hearing loss, but it would be beneficial to provide more data with different degrees of hearing loss and hearing loss at different frequencies. It is our opinion that the ASSR requires more research to better understand optimal test parameters for use as a screening or diagnostic measure for EHDI programs before being clinically implemented. When compared to the ABR and screening measures, ASSR thresholds underperform in their ability to detect *mild* low frequency CHL. The ABR is widely used, with well-studied diagnostic criteria and protocols and it is known that the ABR is accurate in differentiating normal hearing sensitivity from a variety of degrees of CHL, mixed and sensorineural hearing loss in infants. At this time, the ABR continues to be the gold standard and is the diagnostic test method that clinicians should continue to use in their EHDI Programs.

## Limitations and Future Directions

The study was conducted in the context of a clinical audiology department in Canada, where it is routine to have one individual making response judgements. This context was perhaps a limitation of the study. It should be noted that the same individual made threshold estimations for AC and BC ASSRs and ABRs, and hence any subjective bias across response measures should have been similar for all measures, and hence it is unlikely the choice of a single expert judge of response presence is a confounding factor in this study.

The moderate number of infants included in this study demonstrated a mild degree of CHL that was only observed at 500 Hz. The conclusions regarding the efficacy of the ASSR in detecting CHL could differ in a sample with more significant degrees of CHL, hearing loss that extends beyond 500 Hz, and in cases of more extensive middle ear dysfunction (e.g., congenital aural atresia, congenital fixation of the ossicular chain, acute otitis media). The results of the present study suggesting that the ASSR may not be as good an indicator of ABG and CHL (or middle ear abnormality) as the ABR may be limited to ASSR protocols using multitone AM^2^ stimuli, and only for infants with what appears to be a rather mild conductive loss. Further studies with more participants and degrees of CHL may help to provide a larger picture of the efficacy of the ASSR in detection CHL in infants. Of note, acoustic reflexes were not measured in the initial assessment of infants in either NH or CHL groups. In future studies with a wider range of hearing loss degrees, the addition of acoustic reflexes may be beneficial to include to more completely assess in middle ear status. Acoustic reflexes can be problematic, however, in that they may wake the infant and thus are a lower priority measure in most Canadian ABR protocols.

Monotic multiple (MM) ASSR stimuli (500, 1000, 2000, 4000 Hz) were presented; however, due to time constraints only 2000 and 500 Hz were able to be measured down to true threshold and compared with ABR thresholds in this study. The assumption is that if this were to be used clinically as the only electrophysiologic measure of hearing, the clinician would ideally assess threshold to all four stimuli to provide a more complete threshold assessment. In previous studies, it has been demonstrated in human subjects that the use of multiple SAM stimuli is more efficient than a single SAM stimulus (Hatton & Stapells, 2011) and the present study aimed to investigate a possible alternative to the ABR using a technique that more time efficient than the single-frequency technique that is used in diagnostic ABR assessments. In this particular instance, focusing on two frequencies of interest (500 Hz, 2000 Hz) stimuli, while presenting four AM^2^ stimuli may have added some noise to the recordings, and perhaps modestly elevated ASSR thresholds, but the authors are not aware of any published evidence that MM results in higher thresholds than MS in human subjects. It should be noted that four SAM^2^ stimuli were used for both AC and BC ASSR stimuli, and hence would not have expected this to substantially influence the magnitude of the ABG. Future studies aimed to provide clinical evidence of threshold changes resulting from the use of multiple ASSR stimuli would directly address this possibility.

This study aimed to answer a research question while providing some clinically relevant information. As such the test protocol was inefficient and if adopted in a routine clinical setting, would almost certainly have resulted in limited or inadequate information being acquired before the infant wakes. The key procedural elements that would need to be changed in the clinical setting include the test strategy of testing down to true threshold for all infants (clearly unnecessary once it is known that their hearing is ‘Normal’), as well as the use of both the ABR and ASSR as electrophysiological techniques to assess hearing in the same test session (not ever required clinically). The BCEHP protocol for clinical use recommends testing down to a minimum intensity rather than testing down to true threshold in infants with NH (Hatton et al., 2022). In addition, BCEHP does not include the measurement of ASSR thresholds in their clinical protocol; ASSR measures were made solely for the purposes of this study. Using only one electrophysiologic measure of hearing thresholds would significantly reduce test time and would allow for frequency-specific assessment of both ears in a time period that is more reasonable in a clinical setting. An important additional procedural consideration is that ABR testing was the priority for the clinical portion of the assessment, always taking place first. This may have had an impact on the noise of the later-recorded ASSR recordings. It is also worthwhile to explore test time and protocol efficiency of ASSR where there is a consideration for implementation in a clinical setting that is unique to the stimulus and recording parameters and protocols intended to be used (e.g., Cebulla & Stürzebecher, 2015; Sininger et al., 2018, 2020). The difference in starting levels (5-dB offset) between ABR and ASSR may have influenced the differences in ABG size between the two methods. Once correction factors for ASSR are confidently established, a different starting level for ASSR may be more appropriate.

Finally, due to the properties of the underdeveloped skull, the threshold for BC stimuli at 500 Hz is lower (better) in infants than in adults (e.g., Cone-Wesson & Ramirez, 1997; Small & Stapells, 2008a). A logical next step is to measure ABG differences in infants after applying infant-specific correction factors. This study reported ABR and ASSR threshold without the use of any correction factors as these are not yet available for the ASSR (especially for BC stimuli). Future research to establish ASSR eHL correction factors may assist in the clinical application of ASSR thresholds, and act as a spring board for future research involving ASSR ABG in infants with CHL.

## Conclusion

EHDI Programs aim to identify hearing loss early in young infants, with many including mild hearing loss in their target population. Especially outside of North America, there continues to be the perception that obtaining ABR thresholds to low-frequency tone-bursts are too problematic for clinical use. This is not the experience within Canadian EHDI programs (Bagatto, 2020; Hatton et al., 2022), nor of the present study. Tone-evoked ABR for low- to high-frequency stimuli is indeed feasible and can be used to separate mild conductive hearing loss from normal hearing. In contrast, the ASSR may be more problematic. Compared to the ABR, the variability of ASSR thresholds and ABG size was too great to reliably separate normal hearing from mild conductive hearing loss, at least, using the parameters outlined. Furthermore, sensitivity and specificity for identifying CHL was highest for AC ABR threshold measurement compared to screening and ASSR threshold measurements. This finding supports continuation of the current practice using the ABR as the primary tool to assess hearing thresholds in young infants in Canadian EHDI programs. Before considering the ASSR as a diagnostic tool in this context, more research is needed using infants with varying degrees of conductive hearing loss at multiple frequencies to fully assess its appropriateness.

## Data Availability

Data is not available.

## Acknowledgements

Dr. Susan Small was supervisor and mentor for the first author’s M.Sc. thesis research. Sadly, Dr. Small passed away in 2022. Thanks to Dr. Charlotte Douglas for her assistance in obtaining ethics approval, recruitment of participants and supervision of data collection; Drs. Robert Burkard and David Stapells for their mentorship in the preparation of this manuscript, and Alex Gascon for his assistance with data analysis.

## Notes

This research was supported by a Discovery Grant from the Natural Sciences and Engineering Research Council of Canada to Dr. Susan Small. Ethics approval for this project was obtained from the University of British Columbia’s Clinical Research Ethics Board (certificate # H13-02445) and the University of Saskatchewan’s Biomedical Research Ethics Board (certificate # 13-319).

### Competing Interest Statement

The authors have declared no competing interest.

### Funding Statement

This research was supported by a Discovery Grant from the Natural Sciences and Engineering Research Council of Canada to Dr. Susan Small.

### Author Declarations

Ethics committee of the University of British Columbia and the University of Saskatchewan gave ethical approval for this work.

### Summary of Updates

Minor edits to address reviewers' comments.

